# Two founder variants account for over 90% of pathogenic BRCA alleles in Orkney and Shetland

**DOI:** 10.1101/2024.04.03.24305239

**Authors:** Shona M. Kerr, Lucija Klaric, Marisa D. Muckian, Emma Cowan, Lesley Snadden, Gannie Tzoneva, Alan R. Shuldiner, Zosia Miedzybrodzka, James F. Wilson

## Abstract

For breast and ovarian cancer risk assessment in the isolated populations of the Northern Isles of Orkney and Shetland (in Scotland, UK) and their diasporas, quantifying genetically drifted *BRCA1* and *BRCA2* pathogenic variants is important. Two actionable variants in these genes have reached much higher frequencies than in cosmopolitan UK populations. Here, we report a *BRCA2* splice acceptor variant, c.517-2A>G, found in breast and ovarian cancer families from Shetland. We investigated the frequency and origin of this variant in a population-based research cohort of people of Shetland ancestry, VIKING I. The variant segregates with female breast and ovarian cancer in diagnosed cases and is classified as pathogenic. Exome sequence data from 2,108 participants with three or more Shetlandic grandparents in VIKING I was used to estimate the population prevalence of c.517-2A>G in Shetlanders. Nine VIKING I research volunteers carry this variant, on a shared haplotype (carrier frequency 0.4%). This frequency is ∼130-fold higher than in UK Biobank, where the small group of carriers has a different haplotype. Records of birth, marriage and death indicate genealogical linkage of VIKING I carriers to a founder from the Isle of Whalsay, Shetland, similar to our observations for the *BRCA1* founder variant from Westray, Orkney. In total, 93.5% of pathogenic BRCA variant carriers in Northern Isles exomes are accounted for by these two drifted variants. We thus provide the scientific evidence of an opportunity for screening people of Orcadian and Shetlandic origins for each drifted pathogenic variant, particularly women with Westray or Whalsay ancestry.

## Introduction

Pathogenic variants in *BRCA1* and *BRCA2* confer a high lifetime risk of breast, ovarian and, for *BRCA2*, male breast and prostate cancer (1–3). Genetic testing for disease-associated variants in these genes is increasingly available as part of routine clinical care, to enable not only early detection and risk reduction, but also to guide cancer treatment decisions. The genes are considered “tier 1” genes by the Centers for Disease Control and Prevention Office of Public Health Genomics, on the basis that early detection of pathogenic variants will have a positive effect on public health. Predictive testing of unaffected family members is well established, with female pre-symptomatic carriers of pathogenic variants being offered risk-reducing salpingo-oophorectomy and mastectomy, and early onset annual MRI breast screening. Pathogenic variants in *BRCA2* also increase risk of prostate cancer and male breast cancer (4), with male pre-symptomatic carriers typically being offered prostate-specific antigen screening in the NHS. Compellingly, the Icelandic p.Asn257LysfsTer17 founder variant in *BRCA2* (previously termed 999del5 (5)) was associated with a shorter lifespan for both female carriers (effect of the variant, −9.3 years) and male carriers (effect of the variant, −4.6 years) (6).

In isolate populations, a pathogenic variant present in a founding or early member can become more frequent in later generations. Pathogenic variants in *BRCA1* and *BRCA2* that have drifted up in frequency due to founder events have been described in many isolate and founder communities worldwide, notably Ashkenazi and Sephardi Jews (7, 8), and Icelanders (5), as well as in larger populations (9).

The Northern Isles of Scotland – the Orkney and Shetland archipelagos - contain the most isolated of all British and Irish populations, with the highest degree of Norse admixture. The genome-wide distinctiveness revealed at common variants (10) is also reflected in the spectrum of rare and ultra-rare variation: many such variants are enriched in the Shetlandic population (11). This pervasive genetic drift also influences variants of clinical importance. Examples in the Northern Isles include a pathogenic *BRCA1* founder variant that has drifted to high frequency in Orkney (12), and an actionable *KCNH2* Long QT Syndrome variant in Shetland (13). Here, we survey the landscape of pathogenic *BRCA1,* (MIM#113705) *BRCA2* (MIM#600185) and other breast cancer risk variants in the Northern Isles of Scotland.

The NHS Grampian clinical genetics team initially observed the *BRCA2* splice acceptor variant, c.517-2A>G, in breast and ovarian cancer cases from Shetland. The interpretation that this base substitution is a pathogenic variant was demonstrated independently by multifactorial likelihood quantitative analysis (definitely pathogenic, posterior probability 0.9994), together with co-segregation and tumour pathology (14). The c.517-2A>G variant (also known as IVS6-2A>G) has been reported in multiple individuals affected with Hereditary Breast and Ovarian Cancer worldwide, and is classed as pathogenic (reviewed by an expert panel) in ClinVar (15), accession number VCV000051801. This *BRCA2* variant lies in a canonical splice acceptor site and is predicted using a range of tools to affect mRNA splicing, resulting in a significantly altered protein due to exon skipping (16). Experimental evidence *in vitro* supports these bioinformatic predictions (16, 17).

Viking Genes (viking.ed.ac.uk) comprises three cohort studies aiming to explore the genetic causes of disease – the Orkney Complex Disease Study (ORCADES), VIKING I and VIKING II/III. VIKING I contains a rich data resource of more than 2,000 deeply phenotyped and exome-sequenced research subjects with three or four grandparents from Shetland, whereas ORCADES is a similar-sized cohort of participants from Orkney. Most recently, VIKING II/III is a worldwide cohort of 6,000 people, recruited online, with ancestry from the Northern or Western Isles of Scotland (18). The UK Biobank is a cosmopolitan biomedical database containing genetic, lifestyle and health information from half a million participants in the UK (19). Here, we report the ascertainment of the *BRCA2* pathogenic variant c.517-2A>G in both clinical care in cancer families, and research volunteers, from Shetland. We investigate relatedness of the variant carriers in Shetland using genealogy, infer origins, compare haplotypes in VIKING I and UK Biobank carrier participants and estimate population frequencies of pathogenic BRCA variants in Scottish island founder populations both individually and collectively.

## Materials and Methods

### Genotyping

DNA from all VIKING I participants was used for genome-wide genotyping on the GSA BeadChip (Illumina) at the Regeneron Genetics Center. Monomorphic genotypes and genotypes with more than 2% of missingness and Hardy-Weinberg equilibrium (HWE) p<10^-6^ were removed, as well as individuals with more than 3% of missingness. Details of genotyping, sample and variant quality control of UK Biobank genotyping data are described in Bycroft *et al* (20).

### DNA Sequencing and Annotation

The fully quality-controlled exome sequence data set of 2,108 VIKING I sequences (843 male and 1,265 female participants) passed all genotype quality control thresholds as described previously for the exomes in the ORCADES cohort (12). The data was prepared at the Regeneron Genetics Center, following the process detailed for UK Biobank (21). Details of the quality control of whole exome sequencing on the 200,000 participants from the UK Biobank are described in Backman *et al* (22). For Viking Genes, exonic variants passing QC in *BRCA1*, *BRCA2* and other cancer risk genes were filtered against ClinVar (15) (accession date 28th March 2023) and Pathogenic/Likely Pathogenic variants were retained, irrespective of star status. Pre-designed primers (Hs00585575, Thermo Fisher Scientific) were used to generate a PCR fragment of 268 base pairs from genomic DNA for analysis. The *BRCA2* c.517-2A>G, rs81002858 variant was confirmed in VIKING I carriers by Sanger sequencing on an Applied Biosystems 3730xl Genetic Analyser. All nine heterozygous variant calls from the VIKING I exome dataset were verified.

### Haplotype analysis

The VIKING I array genotype data were phased using Shapeit2 v2r837 (23), with the duoHMM option that uses the family-based nature of the data (24). Prior to phasing, the array genotype data were lifted over from the genome build GRCh38 to GRCh37 using LiftOver, followed by quality control against the TOPMED reference panel with Rayner’s HRC-1000G-check-bim (v4.2.13) script that was downloaded from https://www.well.ox.ac.uk/~wrayner/tools/. UK Biobank data were interrogated to ascertain the frequency of the variant of interest. Details of phasing of UK Biobank genotyping data have been described (20). Then, the phased genotypes were used to determine a shared haplotype around c.517-2A>G using the fine method described previously (13). All methods were performed using R 4.0.2 (R Core Team 2020 https://www.R-project.org/). Haplotypes were defined with custom-built in-house scripts in R. Data handling was performed using data.table and tidyverse R packages. Plots were generated using ggplot2.

A single variant-based haplotype search was performed to determine the haplotype length between the different VIKING I carrier kindreds, and with the UK Biobank carrier individuals, using a stepwise approach. Using phased genotype data, starting from the rs81002858 rare variant, one SNP at a time was added to define a haplotype. The procedure was repeated until haplotypes of two individuals (both known carriers) no longer matched, both upstream and downstream of the rare variant, providing variant-level resolution of the haplotype length. The procedure was repeated for all pairs of individuals identified as carriers based on the exome sequencing data, both in the VIKING I and UK Biobank datasets. The shortest shared haplotype from VIKING I was then merged with the shortest shared haplotype from the UK Biobank, to compare whether the haplotypes match.

### EHR Data Linkage in VIKING I

NHS routine datasets linked to VIKING I participants in July 2021, including the Scottish Cancer Registry SMR06, were accessed using a secure process, as described previously (12).

## Results

### Clinical ascertainment of the BRCA2 c.517-2A>G variant

Eleven women from four Shetland families presenting with *BRCA2*-related cancers were found to have the *BRCA2* c.517-2A>G variant, as part of routine clinical care. The family branches share some common surnames, but the branches could not be genealogically linked. Two of the branches overlap genealogically with two VIKING I families with the same variant. To date, five of these 11 clinically ascertained carriers have had breast or ovarian cancer. Not all would have met the criteria for genetic testing in the mainstream NHS setting (see Supplementary Methods), without the context of the wider family health history.

### Population frequencies of the BRCA2 c.517-2A>G variant

Data from 2,108 VIKING I participants passed all exome sequence and genotype quality control thresholds. There are nine heterozygous carriers of the *BRCA2* c.517-2A>G variant in the VIKING I exome dataset, of whom six are female. The Genome Aggregation Database (gnomAD v4.0) (25), containing over half a million exomes from unrelated individuals, has 14 heterozygous carriers of the c.517-2A>G variant (Table 1). Consistent with this rarity, the variant is not observed in 2,090 participants in ORCADES (12). In good agreement with gnomAD, 458,838 whole genome sequences from the UK Biobank (26) contain fifteen instances (Table 1).

**Table 1.** Frequencies of *BRCA2* c.517-2A>G carriers in genomic datasets. . The numbers of carriers are reported for diverse genomic datasets. gnomAD, genome aggregation database; CanVar-UK, Cancer Predisposition Gene Variant Database.

| Dataset | Description | Number of Individuals | Carriers of *BRCA2* c.517-2A>G variant | Non-European Carriers |
| --- | --- | --- | --- | --- |
| gnomAD v4.0 | Unrelated European (non-Finnish) individuals from genetic studies | 549,120 exome sequences | 14 | 0 |
| Viking Health Study Shetland (VIKING I) | Isolate population cohort from Shetland, UK | 2,108 exome sequences | 9 | 0 |
| Orkney Complex Disease Study (ORCADES) | Isolate population cohort from Orkney, UK | 2,090 exome sequences | 0 | 0 |
| UK Biobank | Cosmopolitan population cohort from UK | 458,838 whole genome sequences | 15 | 0 |
| CanVar-UK | Sequenced cancer genes of NHS cancer patients England & Wales | 80,722 total probands tested | 35 | 5 |
| Japanese women with breast cancer | Direct sequencing of *BRCA1* and *BRCA2* genes | 325 women with breast cancer | 1 | 1 |

We have access to exome sequence data from 200,000 individuals from UK Biobank, six of whom carry the variant. These six were born and live in England, mostly in Yorkshire. Variant carriers are also reported in a national UK repository of variants in cancer predisposition genes in cancer patients, CanVar-UK (27). There are 35 carriers in this dataset (Table 1), five of whom are recorded as of non-European heritage. The variant was also noted in one participant in a study of Japanese women with breast cancer (28).

### Origin of the c.517-2A>G variant in Shetland

Three out of nine of the VIKING I variant carriers had all four grandparents born in the Isle of Whalsay, one of the North Isles of Shetland, and five of them had at least one Whalsay grandparent. Of all 36 grandparents of the carriers, 14 (39%) were from Whalsay, with most of the remainder coming from other parishes or isles of Shetland (Figure 1). The *BRCA2* pathogenic variant c.517-2A>G in Shetland thus shows evidence of a founder from the parish of Whalsay.

**Figure 1.**
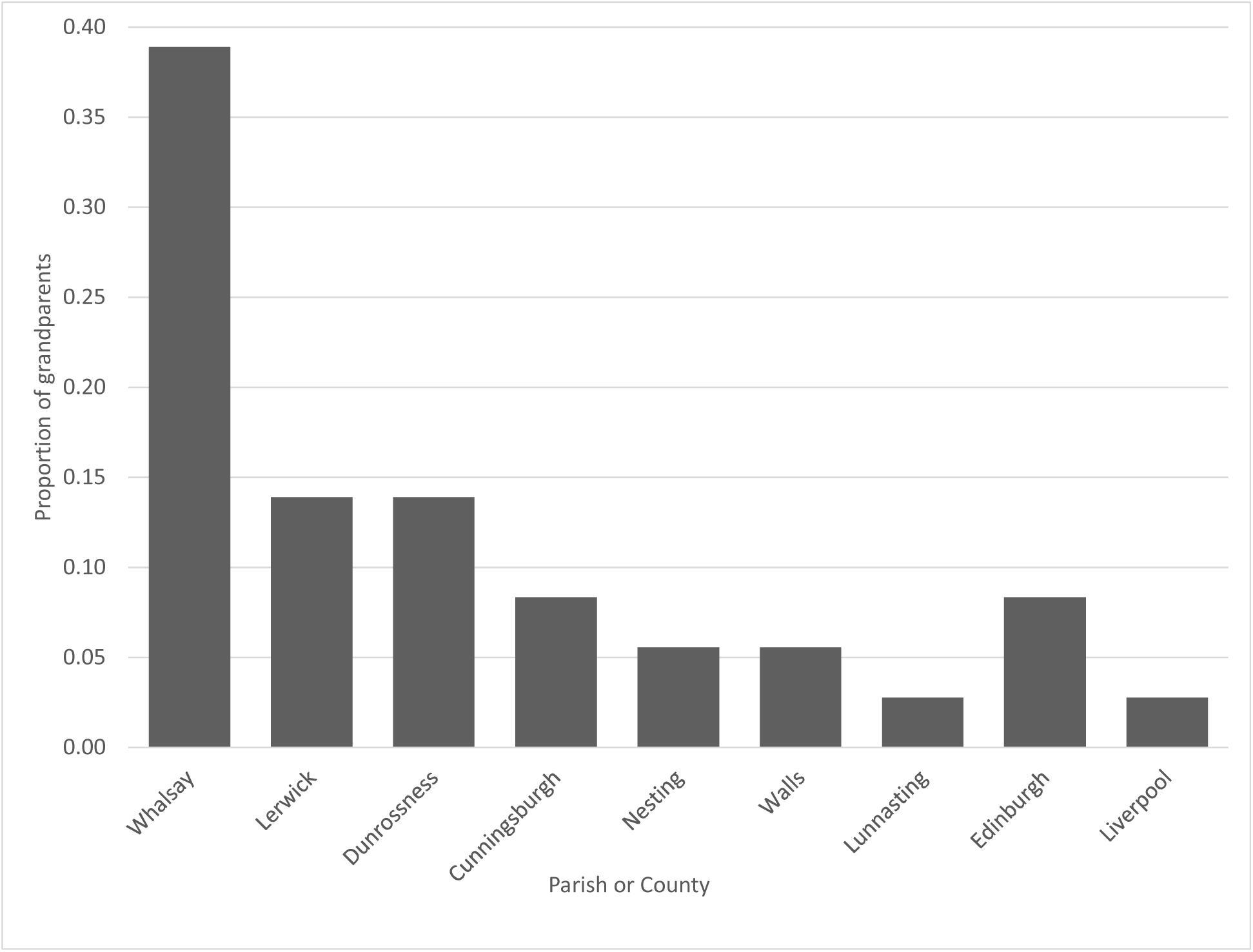
Grandparental ancestry of *BRCA2* c.517-2A>G carriers in VIKING I. The first seven bars are parishes or isles of Shetland. The remainder are locations elsewhere in the UK. Of all 36 grandparents of the carriers, 14 (39%) were from Whalsay, with most of the remainder coming from other parishes or isles of Shetland.

Church records and data from the Scottish register of births, marriages and deaths traced six of the nine research carriers to lineages with an origin in a couple born over 200 years ago on the island of Whalsay, in the Shetland Isles (Figure 2). A further carrier descends from the same ancestral surname in the same isle, potentially linking into the pedigree ∼300 years ago (not shown), but the genealogical records do not survive to prove the link. The c.517-2A>G families in the research cohort can thus be traced back to this family, apart from two closely related carriers who have no genealogical links and no known ancestors from Whalsay. This “Family B” is not shown in Figure 2, and is probably the result of an ancestral non-paternity event, given they share a haplotype with the main kindred and its outlier (see below).

**Figure 2.**
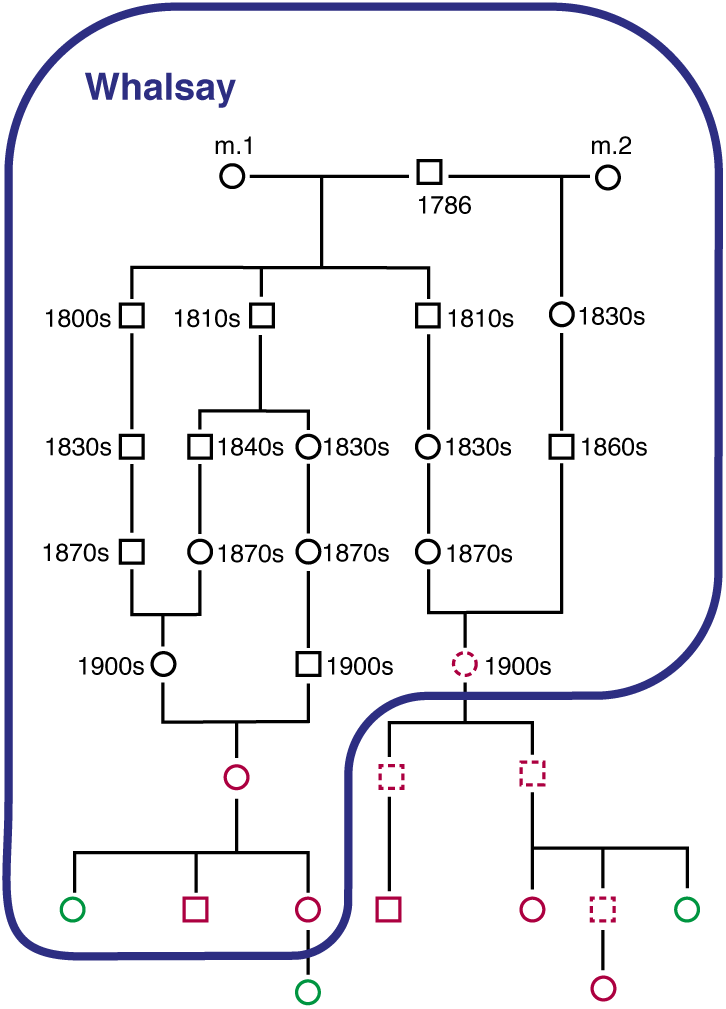
Outline pedigree of a large kindred from the VIKING I study. The founder of the kindred was born in Whalsay in 1786, and all descendants enclosed by the blue line were also born there. He had children by two wives. Red outlines are sequenced *BRCA2* c.517-2A>G carriers in VIKING I, dotted red outlines are obligate carriers (parents of carriers, who link to other carriers). Sequenced VIKING I participants without the variant are shown in green. There are many other family members not shown.

### c.517-2A>G carriers in VIKING I and UK Biobank do not share a common haplotype

All nine heterozygous c.517-2A>G carriers in VIKING I share the same haplotype around the variant locus, with a minimum length of 3.6 Mb (Figure 3). Analysis of haplotypes in the genotype data from six UK Biobank participants carrying the variant showed that all but one share a minimum ∼0.7 Mb haplotype around the variant. However, this was different to the Whalsay haplotype from VIKING I, and is noticeably shorter: on average 1.4 Mb are shared vs. 9 Mb among the Shetlanders (excluding relatives closer than fifth degree). The final UK Biobank carrier shares neither of these haplotypes.

**Figure 3.**
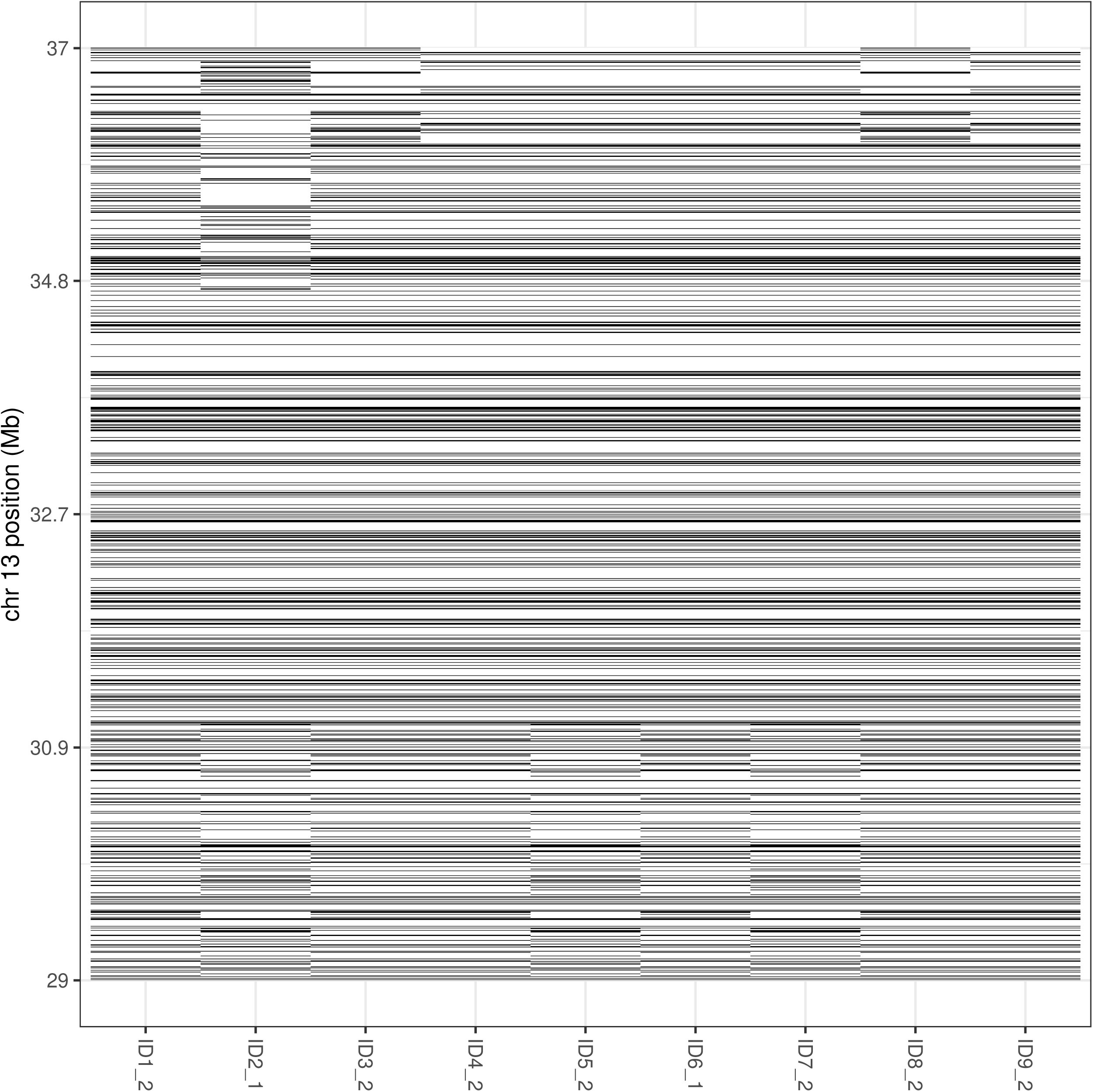
Visualisation of haplotype sharing across nine Shetland carriers. The phased Illumina GSA chip genotypes across an 8 Mb region on chromosome 13 are represented for one haplotype from each individual. The pathogenic variant is at 32.33 Mb and a multi-Mb identical-by-descent block is apparent in the centre of the plot. For close relatives, the extent of sharing goes beyond the window shown. Mb, megabase.

### Penetrance of BRCA2 c.517-2A>G

Linkage to routine NHS data in the electronic health record (EHR) provides a longitudinal component to the Viking Genes research cohorts, and affords the opportunity for unbiased penetrance estimates. All nine c.517-2A>G carriers gave consent for EHR linkage, and the mean age of the six female carriers at time of recruitment to VIKING I (baseline in 2013-2015) was 52. Analysis of the NHS cancer registry (SMR06) dataset shows that one of the female c.517-2A>G carrier participants has a diagnosis of breast cancer, and one of ovarian cancer. None of the other seven carriers (four females and three males) had any cancer code recorded, or codes in the hospital admissions (SMR01) dataset for relevant risk-reducing surgery, and all nine were alive in 2021. Volunteers also complete study questionnaire sections on family history of disease, and this reveals that two obligate carriers – parents of participants, through which they link to other carriers – were affected with cancer. One of these cancers (in a father) was non-Hodgkin’s lymphoma, for which there is some evidence of association with pathogenic variants in *BRCA2* (29).

### The landscape of BRCA1 and BRCA2 pathogenic variants in the Northern Isles

Apart from the two founder variants (*BRCA1* c.5207T>C; *BRCA2* c.517-2A>G), in our ∼4,200-person Northern Isles sample, only four other actionable BRCA variants are observed, all of them singletons in *BRCA2* (Supplementary Table 1). Two of these were inherited by participants from non-Northern Isles parents, meaning that 93.5% of actionable BRCA variant carriers in Northern Isles genomes are accounted for by the two drifted variants. One of the singletons inherited from outside the Isles (from a NE Scottish Mainland-born parent) was the NE Scottish founder variant *BRCA2* c.6275_6276del p.Leu2092fs (also called 6503delTT; rs11571658 (30)). The other noted West Central Scottish founder variant, *BRCA1* 2800delAA (31), was absent from our cohorts.

### Allele frequencies of founder BRCA variants

To explore further how common the drifted c.517-2A>G *BRCA2* variant is across Shetland, we filtered our population for people with four grandparents, and therefore their entire genome, from the Isle of Whalsay. While there will be large confidence intervals, the point estimate of the carrier frequency is 2.3%, or one in 43 people from this isle, five-fold higher than in a general sample of Shetlanders, where the carrier frequency is 1/234 (Table 2). For comparison, we have also analysed data from Orkney for the drifted *BRCA1* c.5207T>C variant, filtering for people with one or more grandparents from the Isle of Westray, Orkney. The genetic drift is more extreme, with a carrier frequency estimate of 5.2% or 1 in 19 Westray genome equivalents, again five-fold higher than in a general sample of Orcadians. These otherwise rare pathogenic alleles are therefore 700-2300 times more common in these isles than in the general UK Biobank population (Table 2). For added context, numbers are also given in Table 2 for the well-known Icelandic *BRCA2* (6) and Ashkenazi Jewish *BRCA1* and *BRCA2* pathogenic alleles (7), with carrier frequencies between 1 in 138 in Iceland and 1 in 41 for the combination of three variants in the Ashkenazi community.

**Table 2.** Carrier frequency estimates of *BRCA* founder variants in the Northern Isles of Scotland and comparison populations. UKB-EU, UK Biobank European-heritage, exome data reported by CanVar-UK v2.2 release date July 2023 (27). Westray estimates use data from ORCADES on n=1461 self-reported Westray-born grandparents, or 365.25 full Westray genome equivalents. Whalsay estimates use VIKING I individuals with all four grandparents born in Whalsay. N, number of carriers. “One in” gives the reciprocal of the carrier frequency, e.g. one in 105 individuals. Allele frequencies are half the carrier frequencies, for variants such as these, where no homozygotes are observed. Uplift presents the fold uplift in frequency over and above that in UK Biobank European-heritage. The total number of individuals for the sum of the three Ashkenazi founder variants is taken as the average of the counts for the three individual variants. *BRCA2* 517-2A>G is rs81002858; *BRCA1* c.5207T>C is rs45553935; *BRCA2* c.771_775 del is rs80359671; *BRCA1* c.68_69del is rs80357914; *BRCA1* c.5266dup is rs80357906; *BRCA2* c.5946del is rs80359550.

| Variant | Population | N | Total | Frequency | % | One in | Uplift | Reference |
| --- | --- | --- | --- | --- | --- | --- | --- | --- |
| **BRCA2**<br>**c.517-2A>G** | UKB-EU | 14 | 432,222 | 3.24E-05 | 0.0032 | 30,873 |  | 27 |
|  | Shetlanders | 9 | 2108 | 0.0043 | **0.43** | **234** | 132 | current study |
|  | Whalsay | 3 | 129 | 0.023 | **2.3** | **43** | 718 | current study |
| **BRCA1**<br>**c.5207T>C** | UKB-EU | 10 | 442,259 | 2.26E-05 | 0.0023 | 44,226 |  | 27 |
|  | Orcadians | 20 | 2090 | 0.0096 | **0.96** | **105** | 423 | 3 |
|  | Westray | 19 | 365.25 | 0.052 | **5.2** | **19** | 2301 | current study |
| **BRCA2**<br>**c.771_775del**<br>**(999del5)** | Icelanders | 421 | 57,933 | 0.0073 | **0.73** | 138 |  | 14 |
| **BRCA1**<br>**c.68_69del**<br>**(185delAG)** | Ashkenazi<br>Jews | 85 | 9371 | 0.0091 | **0.91** | 110 |  | 15 |
| **BRCA1**<br>**c.5266dup**<br>**(5382insC)** | Ashkenazi<br>Jews | 24 | 8867 | 0.0027 | **0.27** | 369 |  | 15 |
| **BRCA2**<br>**c.5946del**<br>**(6174delT)** | Ashkenazi<br>Jews | 119 | 9514 | 0.012 | **1.2** | 80 |  | 15 |
| **3 Ashkenazi**<br>**founder**<br>**variants** | Ashkenazi<br>Jews | 228 | ~9250 | 0.024 | **2.4** | **41** |  | 15 |

The frequency estimates of BRCA variants in the Northern Isles of Scotland are in total somewhat higher than among women in the UK in general. For example, 1/293 women in UK Biobank are carriers of pathogenic / likely pathogenic (with at least 2 star status in ClinVar) or predicted protein-truncating variants in *BRCA1/2* (32). Hence, the proportion carrying a potentially damaging BRCA variant in Orkney (1/105) is more than twice that elsewhere in the UK. However, as shown in Table 2, it is when the individual isle of origin is considered that the carrier frequencies rise an order of magnitude (e.g. 1/43 in Whalsay, 1/19 in Westray), similar to the 1/41 observed for Ashkenazi Jews (three founder variants).

### Other breast cancer susceptibility genes

Variants in the ataxia telangiectasia mutated gene *ATM* can increase the risk of breast cancer, though the gene is not on the ACMG v3.2 list of recommendations for reporting secondary findings in clinical exome and genome sequencing (33). A pathogenic coding variant in *ATM* (c.7271T>G p.Val2424Gly; rs28904921, ClinVar VCV000003023) was described in an Orkney pedigree in 1998 (34), the “core” family. The variant is rare but widespread globally, and case-control studies indicate it is strongly associated (OR∼10) with breast cancer (35, 36). In the ORCADES research cohort, we recovered the same previously reported family (34). Beyond the previously reported three heterozygotes and three homozygotes, we identified one further heterozygote closely related to this family.

Two further *ATM* p.Val2424Gly carriers were found, one is a seventh degree relative of the core family. In total there are 6 alleles out of 4,180 sequenced in ORCADES = 0.14%. For comparison, in the UK Biobank there are 91 alleles out of 917,638 European alleles sequenced = 0.0099%, thus there is a moderate uplift in Orkney (∼15-fold). We know from questionnaire data that a large number of first and second-degree relatives of the carriers in the core family are alive but not in the ORCADES study, some of whom may have volunteered to join the online study VIKING II (18). This may allow a better delineation of the size of the kindred, and more accurate estimation of the frequency and any impact of the variant on breast cancer risk in the Northern Isles of Scotland.

The relatively common loss-of-function c.1100del variant in *CHEK2* (rs555607708, ClinVar VCV000128042) imparts a moderate risk of breast and prostate cancer, even in individuals unselected for family history (37, 38). We observed only four heterozygous carriers of this variant, two from ORCADES and two from VIKING I, giving an estimated minor allele frequency of 0.048% in the Northern Isles. This is five-fold lower than the 0.24% seen in UK Biobank. It is notable that three of these four carriers in the Viking Genes cohorts derive half their genomes from Lowland Scotland, and in one individual we know the allele must have originated from the Scottish Mainland parent. This suggests c.1100del was rare-to-absent in the pre-WWII Northern Isles gene pool (due to genetic drift), but appears today due to post-WWII immigration from the rest of the British Isles. Like *ATM*, *CHEK2* is not on the ACMG3.2 gene list of recommendations for reporting incidental findings (33), but discussions related to the inclusion of further moderate penetrance breast cancer genes, including both *ATM* and *CHEK2*, are ongoing (39); both of the above variants are tested clinically in the UK, according to set guidelines (40).

No pathogenic exonic variants in other genes which confer an increased risk of breast cancer were discovered: *TP53*, *PALB2*, *STK11*, *PTEN* – suggesting these are also essentially absent from the Northern Isles gene pools. Moreover, none of the *BRCA2* c.517-2A>G carriers in VIKING I also carry known pathogenic exonic variants in the cancer susceptibility genes *APC, BRCA1, RET, MAX, TMEM127, BMPR1A, SMAD4, MLH1, MSH2, MSH6, PMS2, MEN1, MUTYH, NF2, SDHD, SDHAF2, SDHC, SDHB, RB1, VHL* or *WT1*.

In the Northern Isles, the effects of random genetic drift over the centuries have thus simplified the landscape of highly penetrant variants of large effect, such that two variants account for almost all of the risk arising from rare pathogenic hereditary breast and ovarian cancer variants, while the impact of all others is negligible. Clearly, high penetrance genes contribute only a proportion of genetic cancer risk, whereas common low penetrance variants identified through genome-wide association studies explain a further component, as of course do environmental exposures.

## Discussion

We recurrently identified the pathogenic *BRCA2* variant c.517-2A>G in the population of Shetland, both in clinical practice and in a population cohort. The variant is found in ∼0.4% of Shetlanders. All carriers in the Shetland cohort VIKING I share the same haplotype background, which is distinct from that observed in English UK Biobank carriers, pointing to recurrent mutation at the site. Genealogical analysis suggests the variant is likely to have originated or arrived in a founder individual from Whalsay, Shetland, over 300 years ago. The presence of the same variant in a Japanese study, and in five non-white individuals in CanVar, provides further evidence that this variant has arisen more than once worldwide.

Individuals not ascertained by cascade testing of relatives from the clinical pedigrees were identified in the population-based sample. In previous work on the *KCNH2* variant p.Gly584Ser (13), which increases risk of long QT syndrome, and on *BRCA1* p.Val1736Ala (12), we similarly identified carriers in our Northern Isles research populations who could not have been ascertained from family-based cascade testing. This highlights the value of research cohorts in uncovering rare clinical variants, which may in turn help planning and implementation of genetic services. Our online cohort study, VIKING II, which has recruited volunteers with Northern Isles grandparents irrespective of domicile (18), has highlighted scientifically for the first time the extent of emigration. Significant Northern Isles diasporas are found in mainland Scotland, former British colonies, particularly New Zealand, Canada and Australia, and also in England, all locations where these alleles are therefore likely to be important for some families.

We show that the frequency estimates of pathogenic BRCA alleles in Orkney and Shetland are somewhat higher than those for women living in other areas of the UK. We have also demonstrated that variation in *ATM* and *CHEK2* does not contribute significantly to breast cancer in Orcadians. However, the overall numbers affected by high penetrance variants will be low, given the modest population sizes. In the most recent census for which data is available (2011), the population of Whalsay was 1,061 and of Westray, 588, while Shetland and Orkney had 23,167 and 21,279 inhabitants, respectively. The size of the extensive diasporas is unknown. Given the combined population of over 64,000 in 1861 and the estimated number of living descendants that would arise in line with European population growth rates in this period, we speculate that the diasporas number in the hundreds of thousands, although the great majority will have inherited only a fraction of their genomes from Northern Isles ancestors. Ongoing isolate breakdown, due to continued immigration to the isles, but also increased within-archipelago migration and mixing, are expected to similarly result in dilution of these high Northern Isles frequencies over time. High penetrance *BRCA1* and *BRCA2* founder pathogenic variants have been described in several populations such as the Ashkenazim, Icelanders, Poles, Norwegians and others (8). Testing for these variants is increasingly well established in both clinical (https://www.nhsjewishbrcaprogramme.org.uk/) and research (41) contexts and cost-effectiveness has been demonstrated in a number of settings. A recent opinion paper addresses the routine failure to identify many monogenic cases of common diseases, including breast and ovarian cancer, in clinical settings (42). It aims to focus attention on universal genetic testing in common diseases for which the recommended clinical management of patients with monogenic forms of disease diverges from standard management, and has evidence for improved outcomes. In a step towards this, all women with a diagnosis of a high-grade serous ovarian carcinoma in Scotland (https://www.sign.ac.uk/media/2010/sign135_oct2022.pdf) and Wales, UK, are now tested for pathogenic *BRCA* variants, without the need for any family history (43, 44). Furthermore, those with breast cancer under the age of 40, and triple negative breast cancer under 60, regardless of family history, are widely tested in the UK (Supplementary Methods). However, the challenges of potential reduced penetrance in those without a known family history, and cost, have generally limited adoption of asymptomatic *BRCA1* and *BRCA2* screening.

Homozygosity for pathogenic *BRCA2* variants causes a rare inherited form of bone marrow-associated acute myeloid leukaemia, solid tumours and developmental anomalies termed Fanconi anaemia. We estimate that the frequency of Fanconi anaemia in people with Whalsay grandparents could be as high as 1/10,000, considerably higher than the average for Europe of 4-7 per million (45). We propose that awareness raising of this unlikely possibility in clinicians serving this population is important.

Single fragment Sanger sequencing for well characterised founder variants is considerably cheaper than panel, whole exome or whole genome testing. Thus, testing efficiency would be likely to be considerably higher in the Northern Isles of Scotland and their diaspora, as we have shown that ∼93% of all actionable variants in BRCA genes are accounted for by only two drifted variants. Furthermore, negligible numbers of variants are segregating at other breast cancer risk genes. This is in sharp contrast with the 424 different pathogenic or loss-of-function *BRCA1/BRCA2* variants found in UK Biobank females, where 369 variants account for 93% of all actionable variants (*pers. comm.* L. Jackson, University of Exeter, from analysis of pathogenic variants as described in (32)). This considerable simplification of the gene pool in genetic isolates, predicted to be an important factor in gene mapping study designs (46), may in fact be more critical when considering genomic medicine screening in these populations. There is a case for offering all Orcadian and Shetlandic women testing for the relevant drifted pathogenic BRCA variant, particularly those with Westray or Whalsay ancestry. A pilot of self-administered saliva-based testing is underway for the *BRCA1* variant in Westray. Given current fiscal constraints and the somewhat lower frequency of the Whalsay variant, the immediate recommendation is that testing for *BRCA2* c.517-2A>G should be offered to those of Whalsay ancestry with a first-degree relative with breast or ovarian cancer, and, to those with an affected second degree relative connected through an unaffected male.

Currently, the consent framework of most research cohorts – including UK Biobank – does not allow the return of results about carrier status of actionable variants (21). However, participants in such studies often wish to receive results, particularly about “actionable” findings. This has stimulated publication of an international policy for returning genomic research results (47), and a practical checklist for return of results from genomic research in the European context (48). Others also recommend the return of results following detection of hereditary breast and ovarian cancer risk to adult population-based biobank participants (47, 49, 50). We have therefore invited ORCADES and VIKING I participants to consent to return of results. In addition, our recent online cohort study VIKING II/III offered members the option of consent to return of selected clinically actionable results at the point of joining, with an acceptance rate of 98% (18). Future research will explore additional genetically drifted loci in Orkney, Shetland and the Western Isles of Scotland in the Viking Genes research cohorts, implement the return of results to consented participants about actionable variants such as the drifted BRCA variants described here, and evaluate the implementation of saliva-based BRCA testing of DNA in those of Westray and Whalsay origins.

## Data Availability Statement

There is neither Research Ethics Committee approval, nor consent from participants, to permit open release of the individual level research data underlying this study. The datasets generated and analysed during the current study are therefore not publicly available. Instead, the research data and/or DNA samples are available from on reasonable request, via managed access, following approval by the Data Access Committee and in line with the consent given by participants.

## Data Availability

All research data produced in the present study are available from on reasonable request, via managed access, following approval by the Data Access Committee and in line with the consent given by participants.

## Acknowledgements

The study team wish to thank staff from the NHS Grampian genetics team and the Viking Genes Study for their contribution to these datasets. DNA extractions were performed at the Edinburgh Clinical Research Facility, University of Edinburgh. Sanger sequencing was performed by Camilla Drake and the technical services team at the MRC HGU. Emily Weiss and Reka Nagy assembled the ORCADES pedigree, and Barbara Gray the Shetland pedigree, using records at the General Register Office and study information, building on earlier pedigree work by Ruth McQuillan and Jim Wilson. We thank Thibaud Boutin for phasing the GSA chip data and Kiera Johnston for help with analysis of other cancer susceptibility genes. The data in the EHR was provided by patients and collected by the NHS as part of their care and support. We would also like to acknowledge the invaluable contributions of the research nurses in Orkney and Shetland and the administrative team in Edinburgh, and the NHS Grampian genetic counselling team for counselling of the carriers identified in the cohorts. Finally, and most importantly, we thank the people of the Northern Isles for their involvement in and ongoing support for our research.

## Author Contribution Statement

SK managed the project and drafted the manuscript. LK analysed the exome datasets. MM and LK did the haplotype comparisons. EC and LS provided de-identified clinical data. GT and ARS conceived and managed the Viking Genes exome sequencing. ZM recognised carrier patients through the NHS Grampian clinical genetics service, led clinical aspects of return of results planning and proposed policy. JFW is the Chief Investigator of Viking Genes, was awarded funding to implement the work, did genealogical analyses, interpreted the data and helped draft the manuscript. All authors provided input and feedback on drafts of the manuscript.

## Funding

This work was funded by the MRC University Unit award to the MRC Human Genetics Unit, University of Edinburgh, MC_UU_00007/10 and a Wellcome Trust Institutional Translational Partnership Award (University of Edinburgh 222060/Z/20/Z -PIII031). LK was supported by an RCUK Innovation Fellowship from the National Productivity Investment Fund (MR/R026408/1).

## Ethical Approval

Eligible participants were recruited to ORCADES and the Viking Health Study Shetland (VIKING I), now part of Viking Genes, Research Ethics Committee reference 19/SS/0104. Research participants gave written informed consent for research procedures that included electronic health record linkage and DNA sequencing. The data linkage and access to NHS Scotland-originated data for ORCADES and VIKING I was approved by the Public Benefit and Privacy Panel for Health and Social Care (Ref 1718-0380). This research has also been conducted using data from UK Biobank, as part of project number 19655. Frequencies for the *ATM* and *CHEK2* variants reported here, and the *BRCA2* variant c.517-2A>G numbers in Table 1, were derived from the UK Biobank Whole Genome Sequencing (WGS) project and were obtained from the UK Biobank Allele Frequency Browser (afb.ukbiobank.ac.uk), which was generated by the WGS consortium under the UK Biobank Resource (project ID 52293).

## Competing Interests

AS and GT are employees and/or stockholders of Regeneron Genetics Center or Regeneron Pharmaceuticals.

For the purpose of open access, the author has applied a Creative Commons Attribution (CC BY) licence to any Author Accepted Manuscript version arising from this submission.

---

**Supplementary Table 1 Actionable BRCA1/2 variants in Orkney and Shetland.** Pathogenic or likely pathogenic variants in *BRCA1* or *BRCA2* among 4198 exomes from the Northern Isles. All six variants have 3* status in ClinVar as pathogenic and were reviewed by an expert panel, all are autosomal dominant. c. gives the coding DNA position and change; p, the protein position and change; rsid, reference SNP cluster ID; GRCh38, Genome Reference Consortium human build 38; LOF, loss of function.

## Notes

### Funding Statement

This study was funded by the MRC University Unit award to the MRC Human Genetics Unit, University of Edinburgh, MC_UU_00007/10 and a Wellcome Trust Institutional Translational Partnership Award (University of Edinburgh 222060/Z/20/Z -PIII031). LK was supported by an RCUK Innovation Fellowship from the National Productivity Investment Fund (MR/R026408/1).

### Author Declarations

The South East Scotland Research Ethics Committee of NHS Lothian gave ethical approval for this work, reference 19/SS/0104.

