## Supplementary Methods for "Two founder variants account for over 90% of pathogenic BRCA alleles in Orkney and Shetland"

*Research Volunteer* *Recruitment*

Recruitment to ORCADES (Orkney Complex Disease Study) was from 2005-2011 (1) and to VIKING I (Viking Health Study Shetland) from 2013-2015 (2). Blood (or very occasionally, saliva) samples from participants were collected, processed and stored using standard operating procedures and managed through a laboratory information management system at the Edinburgh Clinical Research Facility, University of Edinburgh.

*Cohort Pedigree Information*

Records of the births, marriages and deaths in Orkney and Shetland are held at the General Register Office for Scotland (New Register House, Edinburgh). These records, along with Church records, relationship information obtained from study participants, the Shetland and Orkney Family History Societies and genealogies available online, were used to construct extended pedigrees of study participants in RootsMagic software (S&N Genealogy Supplies), which were then amended to reflect the genetic kinship between individuals using genotype data.

*NHS Testing Clinical Guidelines*

In the UK NHS, germline BRCA1/2 cancer gene testing at diagnosis is offered to:

- all women with breast cancer diagnosed at age 40 and under

- all women with triple negative breast cancer under aged 60 (triple negative means no oestrogen, progesterone or HER2 protein receptors in cancer cells)

- Women with triple negative breast cancer over 60 with a family history of breast or ovarian cancer

- all women with bilateral breast cancer, or two primary breast cancers if average age of diagnoses is under age 60

- anyone with a history of breast / ovarian cancer where the family Manchester score (3) is over 15

- all women with ovarian cancer at any age

- men with breast cancer

Further details are available at <https://www.nice.org.uk/guidance/cg164>
